# Identification of an increased lifetime risk of major adverse cardiovascular events in UK Biobank participants with scoliosis

**DOI:** 10.1101/2022.11.21.22282578

**Authors:** Valentina Q. Santofimio, Adam Clement, Declan P. O’Regan, James S. Ware, Kathryn A. McGurk

## Abstract

**Background:** Structural changes caused by spinal curvature may impact the organs within the thoracic cage, including the heart. Cardiac abnormalities in idiopathic scoliosis patients are often studied post-corrective surgery or secondary to diseases. To investigate cardiac structure, function, and outcomes in participants with scoliosis, phenotype and imaging data of the UK Biobank (UKB) adult population cohort was analysed.

**Methods:** Hospital episode statistics of 502,324 adults were analysed to identify participants with scoliosis. Summary 2D cardiac phenotypes from 39,559 cardiac magnetic resonance imaging (CMR) scans were analysed alongside a 3D surface-to-surface (S2S) analysis.

**Results:** A total of 4,095 (0.8%, 1 in 120) UKB participants were identified to have all-cause scoliosis. These participants had increased lifetime risk of major adverse cardiac events (MACE) (HR=1.45, P<0.001), driven by heart failure (HR=1.58, P<0.001) and atrial fibrillation (HR=1.54, P<0.001). Increased radial and decreased longitudinal peak diastolic strain rates were identified in participants with scoliosis (+0.29, P_adj_<0.05; −0.25, P_adj_<0.05; respectively). Cardiac compression of the top and bottom of the heart and decompression of the sides was observed through S2S analysis. Additionally, associations between scoliosis and older age, female sex, heart failure, valve disease, hypercholesterolemia, hypertension, and decreased enrolment for CMR, were identified.

**Conclusion:** The spinal curvature observed in participants with scoliosis alters the movement of the heart. The association with increased MACE may have clinical implications for whether to undertake surgical correction. This work identifies, in an adult population, evidence for altered cardiac function and increased lifetime risk of MACE in participants with scoliosis. Future genetic analyses would aid to assess causality.

## Introduction

Scoliosis is the lateral curvature of the spine with a Cobb angle >10°, primarily diagnosed in adolescents[1]. There are multiple aetiologies of scoliosis including neuromuscular, congenital, syndromic, or secondary to other diseases such as muscular dystrophy or Friedreich’s ataxia[2]. However, the most common type is idiopathic scoliosis, with a prevalence of 8% in adults aged over 40 years old[3–5]. In later life, scoliosis can result from skeletal muscle diseases such as sarcopenia. Sarcopenia is the loss of muscle mass associated with age that may cause imbalance and alteration on the supportive muscles of the spine, contributing to the progression of degenerative scoliosis in elderly patients[6,7]. Degenerative scoliosis is observed in 68% of adults aged over 60 years old, as the joints and disks of the spine begin to deteriorate[7]. Osteopenia, loss of bone density, is more frequently observed in females and contributes to the severity of the curvature of the spine[8]. Furthermore, the coexistence of congenital heart disease (CHD) and spinal curvature is found in up to 12% of infant and juvenile scoliosis patients presenting with CHD[9,10]. This is likely due to shared developmental aetiology, whereas studies are lacking on the impact of age-related scoliosis on the functioning of a developmentally normal heart.

In addition to impacting physical day-to-day activities, structural disruption of the thoracic cage (pectus deformity) can influence the organs within, such as the heart[11]. Pectus deformities lead to the displacement of the heart towards the left side of the body which can result in right-sided spinal curvatures as the beating heart pushes the thoracic vertebrae to the right[7,11]. Furthermore, the degree of curvature of the spine can greatly increase the risk of restrictive lung disease, which in conjunction with intrathoracic organ displacement, increases the risk of comorbidities with high mortality rates, such as right heart failure[3,12].

The impact of scoliosis on adult cardiac function has not been extensively studied and the relationship between scoliosis and non-congenital cardiac manifestations is not well characterised. In the UK Biobank adult population cohort, we explored whether all-cause scoliosis has an impact on or relationship with cardiac phenotypes of the matured adult heart. We identify altered radial and longitudinal peak diastolic strain rates and an increased lifetime risk of major adverse cardiovascular effects (MACE) in participants with scoliosis in the UK Biobank. In addition, we observe associations between scoliosis and older age, female sex, heart failure, valve disease, hypercholesterolemia, diagnosis of hypertension, and decreased enrolment for cardiac magnetic resonance imaging.

## Methods

### The UK Biobank population cohort

The UK Biobank (UKB) recruited over 500,000 participants aged 40-69 years across the UK between 2006-2010 (National Research Ethics Service, 11/NW/0382, 21/NW/0157)[13]. This project was conducted under the UK Biobank applications 47602 and 40616. All participants provided written informed consent[14].

### UK Biobank codes for identification of scoliosis

A diagnosis of scoliosis was identified for UKB participants through the first occurrence of scoliosis trait (M41*), a composite trait of the first reported date derived by the UKB that incorporates data from primary care, hospital inpatient admissions, death records, and self-reported medical conditions (**Table 2**).

### Cardiac magnetic resonance imaging data

Amongst all UKB participants, 39,559 participants had cardiac MRI (CMR) data available[15]. Imaging was performed using a 1.5 Tesla machines (MAGNETOM Aera, Siemens Healthcare)[15], and 2D summary CMR traits were analysed for an association with scoliosis. This includes left ventricular ejection fraction (LVEF), left ventricular end systolic volume (LVESV), left ventricular end diastolic volume (LVEDV), and measures of cardiac strain: Eulerian radial strain (Err), Eulerian longitudinal strain (EII), Eulerian circumferential strain (Ecc), radial peak diastolic strain rate (PDSR_rr_), and longitudinal peak diastolic strain rate (PDRS_ll_). All CMR traits were adjusted for age at the time of imaging, sex, White British ancestry, systolic blood pressure (SBP), and body surface area (BSA).

### Surface to surface analysis

A mass univariate regression was utilised to explore associations between the three-dimensional (3D) mesh-derived phenotype and scoliosis[16–19]. The underlying principle of this approach is the implementation of a linear regression at each vertex of the 3D atlas to derive a regression coefficient associated with the variable of interest, which results in a map of beta-coefficients showing the strength and direction of these associations. The analysis was adjusted for age at the time of imaging, sex, White British ancestry, BSA, diastolic blood pressure (DBP), and SBP.

### Lifetime risk and survival analyses

Lifetime risk of disease, from birth month to January 2021, was assessed using the first occurrence of scoliosis trait (M41*). The first occurrence of health outcomes summary data in the UKB is reported using ICD10 codes[13]. The fields analysed for the major adverse cardiovascular events (MACE) composite trait were as follows: cardiac arrest, I46*; atrial fibrillation and flutter/arrhythmia: I48* and I49*; heart failure, I50*; and stroke, I64*, as previously published[20]. The survival analysis was conducted using MACE and death as the primary outcome. *Survival* and *survminer* R packages were used to estimate hazard ratios (HR).

### Statistical analysis

R programming language (version 3.6.0) and RStudio software (version 1.3.1073) were used for analyses. Categorical variables were assessed using Chi-Squared Test or Fisher’s Exact Test and expressed as percentages. Continuous variables were assessed using Student’s t-test and expressed as mean ± standard deviation (SD). All P-values were adjusted using Bonferroni correction for multiple comparisons where P_adj_<0.05 was deemed significant.

In the association of disease analysis, all variables were adjusted for age at recruitment (UKB ID: 21022-0.0) and genetic data-derived sex (UKB ID:22001-0.0) using a multiple linear regression model. A separate multiple linear regression model for CMR traits was used to adjust for covariates at the time of imaging: age, sex, White British ancestry, SBP, and BSA.

## Results

### Prevalence of scoliosis in the UK Biobank

The prevalence of all-cause scoliosis in 502,324 participants of the UKB was 1 in 120 (n=4,095; 0.8%). Of the 4,095 participants with scoliosis, 1,489 (37%) were diagnosed with scoliosis prior recruitment. From the total participants with scoliosis, 10 (0.2%) reported congenital scoliosis, 23 (0.6%) reported childhood scoliosis (infantile and juvenile) and the rest reported scoliosis due to other causes later in life. The most common scoliosis subtype was unspecified scoliosis reported by 2,685 (65.6%) participants (**Table 2**).

Participants with scoliosis were significantly older than the rest of the population (59.4 years old ± 7.58; mean age 56.5 years old ± 8.10; P_adj_<0.01) and were more female (69% female in scoliosis cohort; 55% female in the rest of the UKB population; P_adj_<0.01). Lifetime risk analysis showed that participants with scoliosis had significantly longer lives when compared to the rest of the population, regardless of sex (**Figure 1**).

**Figure 1.**
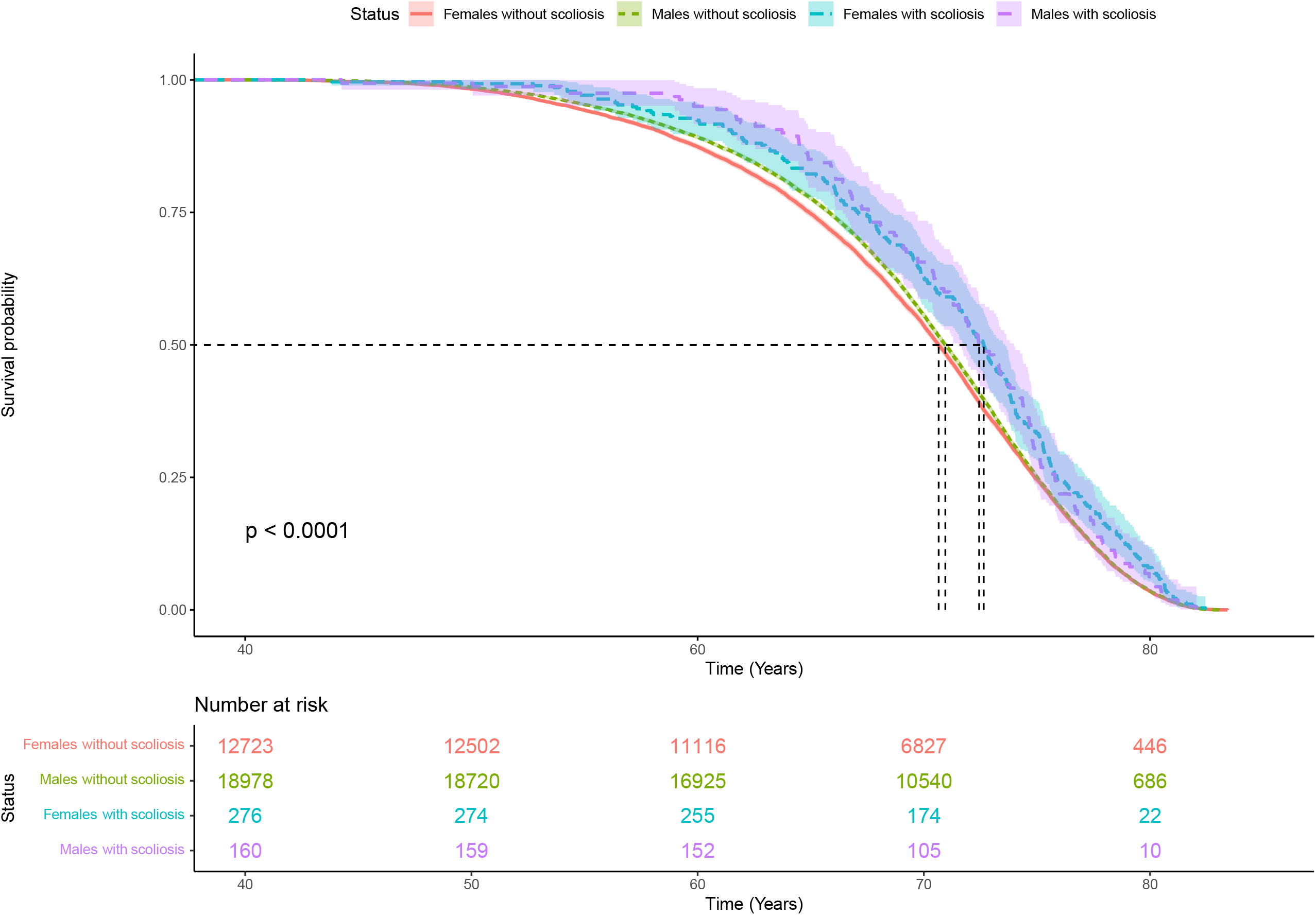
Scoliosis is associated with longer life compared to the rest of the UKB cohort, adjusted for sex. Years of life is shown on the x-axis from date of birth. Counts of UKB participants are shown in the box depicting number at risk. The median survival time (a survival probability of 0.5) was significantly increased in participants with scoliosis, regardless of sex. Median survival time for females with scoliosis vs females without scoliosis (2 years difference); median survival time for males with scoliosis vs males without scoliosis (1.5 years difference).

We observed a significantly increased burden of heart failure (+4%, P_adj_<0.001), valve disease (+1%, P_adj_<0.001), hypercholesterolemia (+7%, P_adj_<0.001) and diagnosis of hypertension (+13%, P_adj_<0.001) in scoliosis participants, adjusted for age and sex (**Table 2**). Although diagnosis of hypertension was significantly increased, no significant differences were found with SBP or DBP, which may be due to correction of diagnosed hypertension through medication. Additionally, the proportion of scoliosis participants with CMR available was significantly decreased (−3%, P_adj_<0.001) compared to the rest of the population.

### CMR analysis of participants with scoliosis identifies altered cardiac diastolic strain

2D summary CMR traits were available on 39,559 participants[15]. 224 participants with scoliosis had CMR available and had significantly increased radial PDSR (P_adj_<0.05) and decreased longitudinal PDSR (P_adj_<0.05) compared to participants without a diagnosis of scoliosis (**Table 3**). Adequate diastolic function is essential during ventricular filling and maintenance of optimum stroke volume. PDSR is a diastolic function trait, that has been previously associated with major adverse cardiovascular events, increased mortality, increased blood pressure and altered left atrial function [21]. For example, decreased PDRS_rr_ corresponds to stiffer ventricle, impairing its relaxation and increasing the risk of heart failure[21]. No additional significant associations were found with the other CMR measures available for analysis and no association was observed between scoliosis and diabetes (**Table 1**).

**Table 1.** Summary statistics of participants with scoliosis compared to the rest of the UK Biobank population cohort. Data is presented as mean ± SD or count (%); all the variables are adjusted for age and sex; BMI, body mass index; SBP, systolic blood pressure; DBP, diastolic blood pressure; CMR, cardiac magnetic resonance imaging; CM, cardiomyopathy. White British ancestry was genetically inferred. P_adj_, P-value adjusted for multiple comparisons via Bonferroni correction; Significant, P_adj_<0.05; NS, not significant, P_adj_>0.05.

| Characteristic | General population<br>(n=502,324) | Scoliosis carriers<br>(n=4,095) | $P_{adj}$ |
| --- | --- | --- | --- |
| Age at recruitment | 56.50 ± 8.10 | 59.40 ± 7.58 | <0.001 |
| White British | 406,131 (81%) | 3344 (82%) | NS |
| Sex, female | 261,862 (55%) | 2,845 (69%) | <0.001 |
| BMI | 27.40 ± 4.80 | 27.00 ± 5.16 | NS |
| Auto SBP | 140.00 ± 19.60 | 141.00 ± 20.00 | NS |
| Auto DBP | 82.20 ± 10.70 | 81.50 ± 10.80 | NS |
| Diabetes | 27,771 (5%) | 212 (5%) | NS |
| Hypercholesterolaemia | 92,824 (18%) | 1,022 (25%) | <0.001 |
| Hypertension | 167,817 (33%) | 1,902 (46%) | <0.001 |
| Included in UKBB CMR cohort | 39,319 (8%) | 224 (5%) | <0.001 |
| Heart failure | 10,504 (2%) | 237 (6%) | <0.001 |
| Any CM | 3,602 (0.7%) | 49 (2%) | NS |
| Valve disease | 5,145 (1%) | 96 (2%) | <0.001 |

**Table 2.** Codes used for identification of all-cause scoliosis. The HES data is coded using the International statistical Classification of Disease (ICD) codes, versions 9 and 10. The self-reported data (SR) has a UKB-specific coding system. SR, non-cancer illness code, self-reported data; ICD9, summary diagnoses (main and secondary); ICD10, underlying primary cause of death, contributory cause of death, and external cause of death, summary diagnoses (main and secondary); GP, general practitioner.

| Disease/phenotype | Participants<br>(n=4,095) | Coding system |
| --- | --- | --- |
| **Self-reported scoliosis** | 468 (11.4%) | SR: 1535 |
| **Kyphoscoliosis and scoliosis** | 5 (0.1%) | ICD9: 7373, 75420 |
| **Congenital postural scoliosis** | 0 (%) |  |
| **Congenital scoliosis** | 10 (0.2%) | ICD10: Q763, M965, M41, M410, M4100-109, M411, M4110-119, M412, M4120-129, M413, M410-139, M414, M1410-149, M415, M1450-159, M418, M4180-189, M419, M4190-199 |
| **Post-radiation scoliosis** | 10 (0.2%) |  |
| **Childhood scoliosis (infantile and juvenile)** | 23 (0.6%) |  |
| **Other idiopathic scoliosis** | 56 (1.4%) |  |
| **Thoracogenic scoliosis** | 24 (0.6%) |  |
| **Neuromuscular scoliosis** | 22 (0.6%) |  |
| **Secondary scoliosis** | 79 (2%) |  |
| **Other forms of scoliosis** | 226 (5.5%) |  |
| **Unspecified scoliosis** | 2685 (65.5%) |  |
| **Unspecified scoliosis** | 487 (11.9%) | GP-derived |

**Table 3.** Summary of the altered cardiac PDSR in participants of the UKB with scoliosis. Mean and standard deviation for adjusted PDSRs are described for the general UKB population and participants with all-cause scoliosis. The p-value was adjusted for multiple comparisons of CMR traits. WT, wall thickness; LVEDV, left ventricular end-diastolic volume; LVEF, left ventricular ejection fraction; LVM, left ventricular mass; Ecc, circumferential; Err, radial; Ell, longitudinal; PDSR_ll_, longitudinal peak diastolic strain rate; PDSR_rr_, radial peak diastolic strain rate.

| CMR trait | General population<br>(n=38,319) | Scoliosis<br>(n=224) | P<sub>adj</sub> |
| --- | --- | --- | --- |
| Maximum WT | 9.40 ± 1.62 | 9.36 ± 1.70 | NS |
| LVEDV | 148.00 ± 33.90 | 136.00 ± 32.80 | NS |
| LVEF | 59.50 ± 6.16 | 59.80 ± 6.03 | NS |
| LVM | 86.0 ± 22.30 | 78.00 ± 20.70 | NS |
| Ecc Global | (-22.33) ± 3.35 | (-22.10) ± 3.77 | NS |
| Err Global | 45.00 ± 8.31 | 45.20 ± 8.73 | NS |
| Ell Global | (-18.50) ± 2.76 | (-17.90 ) ± 2.44 | NS |
| PDSR<sub>ll</sub> | 1.66 ± 0.61 | 1.45 ± 0.58 | <0.05 |
| PDSR<sub>rr</sub> | -5.64 ± 2.09 | -5.08 ± 2.13 | <0.05 |

### Surface to surface analysis of participants with scoliosis shows increased cardiac compression

A 3D surface-to-surface analysis was performed on 21,088 participants of the cohort. The three-dimensional cardiac modelling of scoliosis patients showed increased strain at the top and bottom of the heart (**Figure 2**). Radial cardiac decompression (sides of the heart) was also observed in participants with scoliosis. However, these were not significant when adjusting for the number of comparisons and covariates.

**Figure 2.**
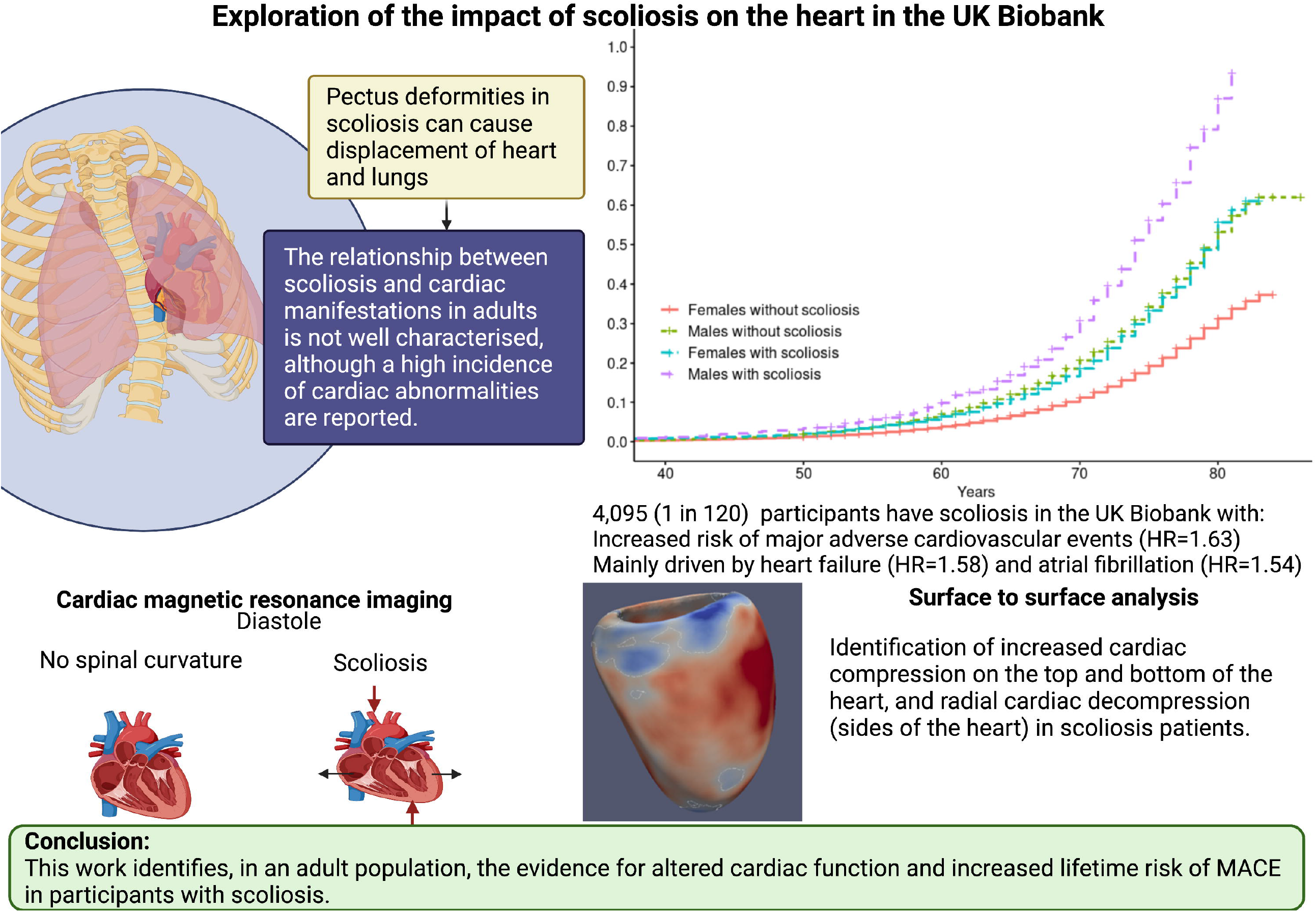
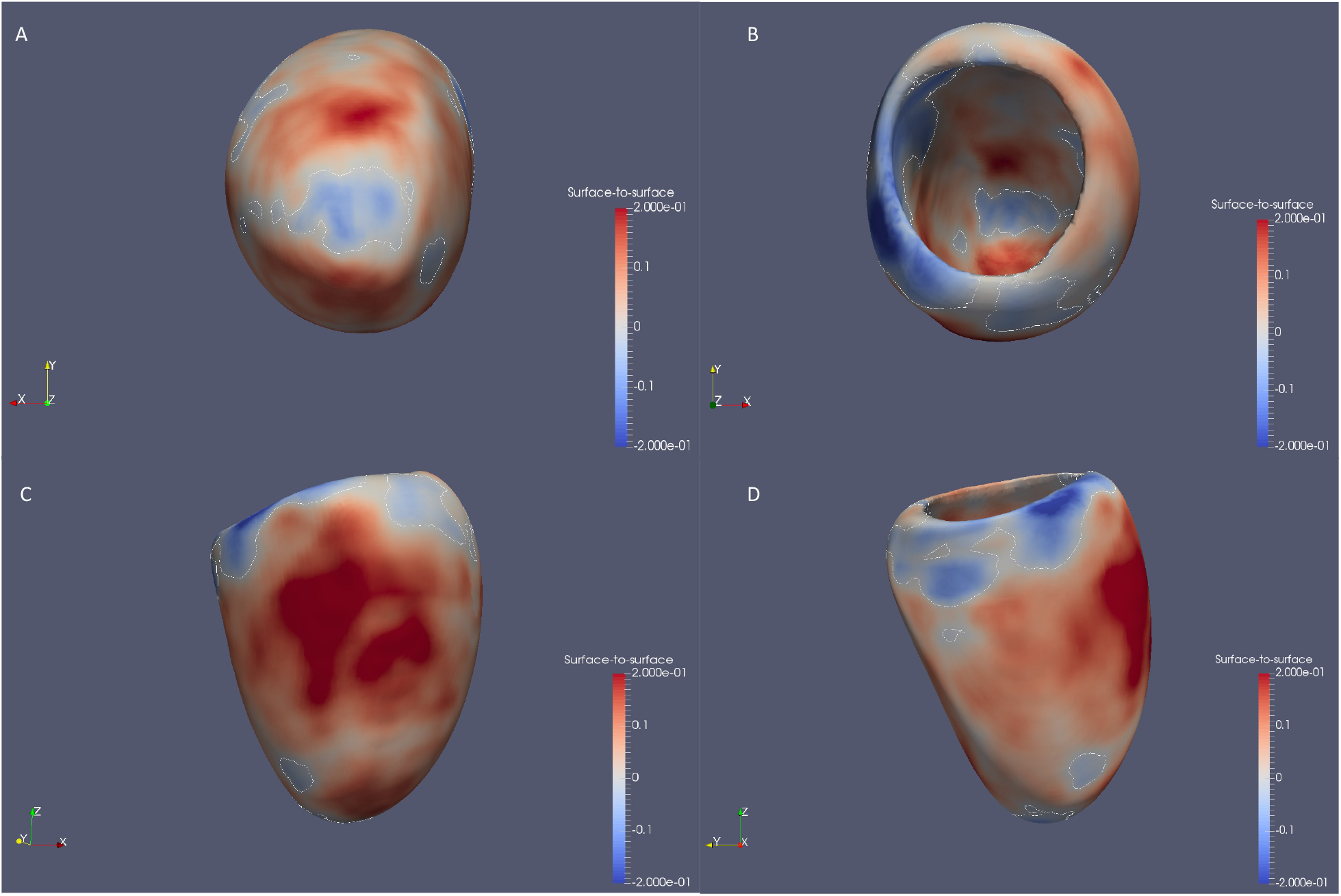
Surface to surface analysis suggests a compressed heart in participants with scoliosis. 3D models of left ventricle geometry with standardized beta-coefficients which show the association between scoliosis and regional surface-to-surface distance. Blue, increased inward pressure; red, increased outwards pressure.

### Lifetime risk of MACE is increased in participants with scoliosis

A significantly increased lifetime risk of MACE was observed for UKB participants with scoliosis (**Figure 3**; HR=1.45, P<0.001), mainly driven by heart failure (HR=1.58, P<0.001) and atrial fibrillation (HR=1.54, P<0.001). The probability of MACE doubled in males into older age (from 60 years of age). This may be caused through the altered cardiac diastolic strain rates observed in participants with scoliosis. However, we emphasise caution regarding the causality of this association of scoliosis and MACE at this stage.

**Figure 3.**
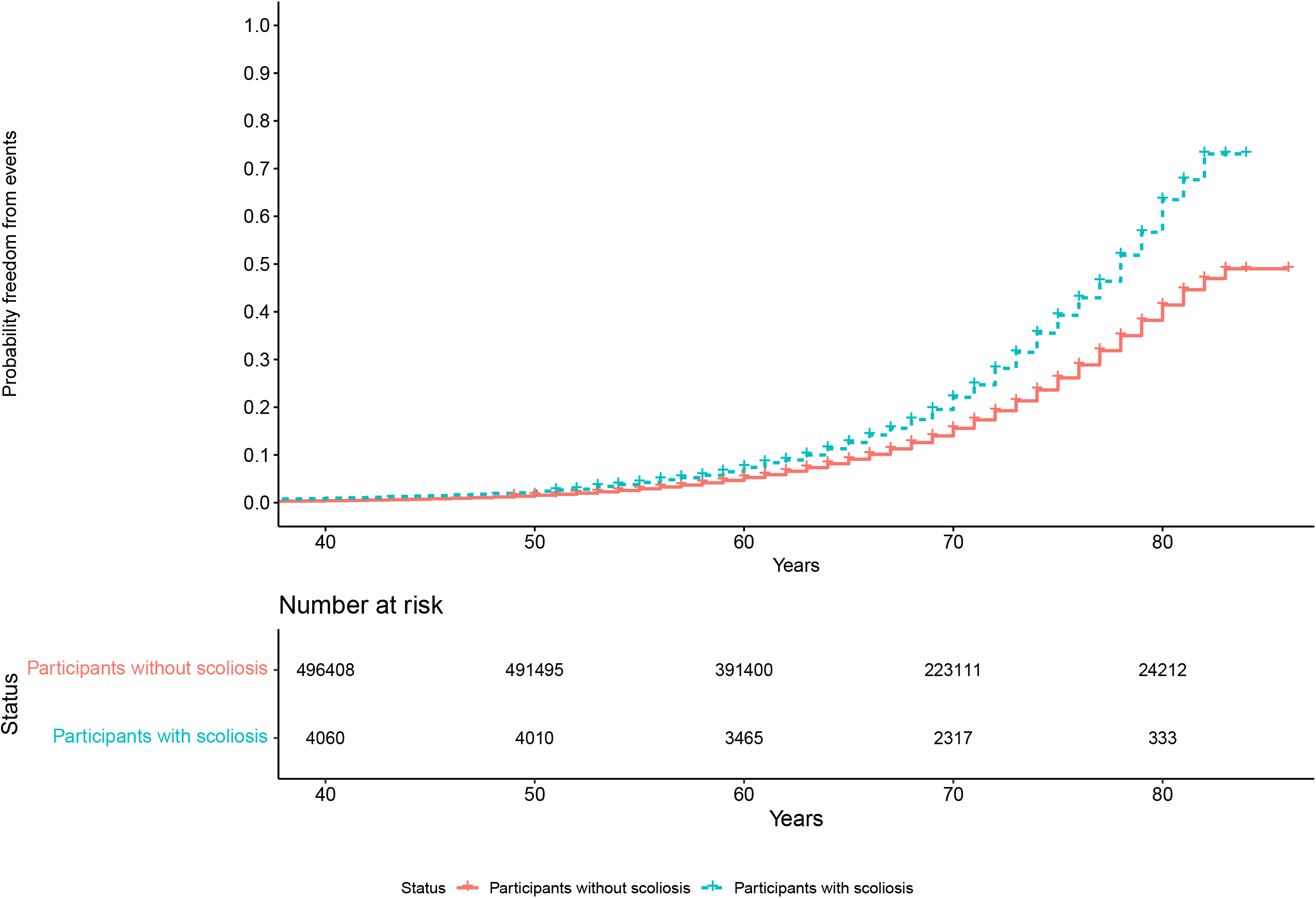
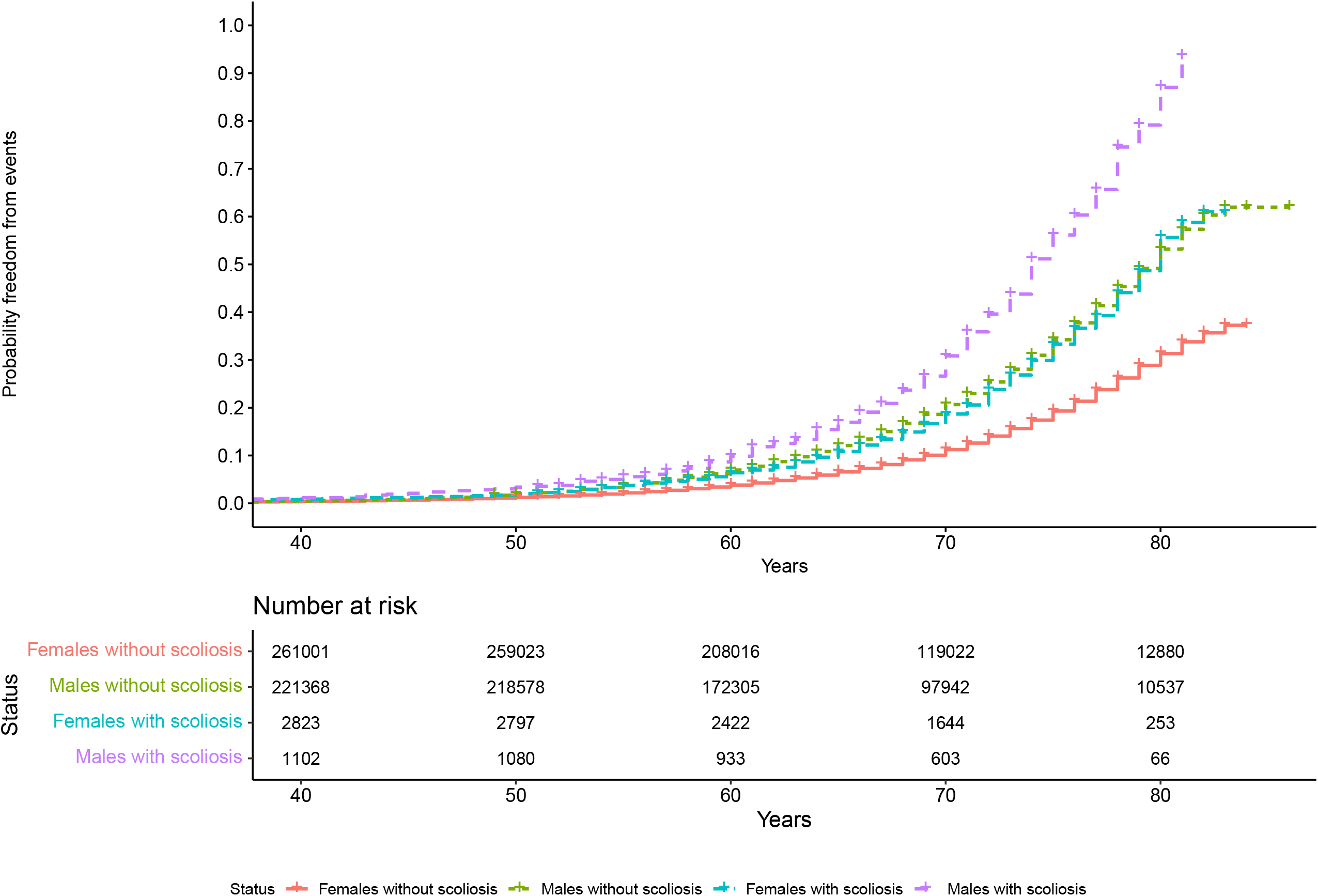
Increased lifetime risk of MACE in UKB participants with scoliosis. A, cumulative incidence curve depicts an increase in lifetime risk of MACE in participants with scoliosis over time (HR=1.45, P<0.001). B, incidence curve stratified by sex (HR=1.63, P<0.001).

## Discussion

### Participants with scoliosis have increased lifetime risk of MACE

To the best of our knowledge, this is the first study to identify an increased lifetime risk of MACE, through atrial fibrillation and heart failure, in participants with scoliosis. This may be through the identified, significantly increased PDSR_rr_ and decreased PDSR_ll_. The abnormal curvature of the spine can increase mechanical constraint on the heart which may result in diastolic dysfunction[22] and the severity of the spinal deformity has been shown to aggravate ventricular and right atrial pressure[3].

It is unlikely that the observed heart failure is due to blood pressure abnormalities as no significant differences were found in measured SBP and DBP between participants with scoliosis and the rest of the population. However, it is possible that pulmonary dysfunction may be eliciting cardiac dysfunction alongside atrial fibrillation, in patients with advanced scoliosis[3,11,23]. These findings suggest that early medical intervention in patients with scoliosis through surgery, may decrease the risk of a future MACE, however future validation of this finding would need to be confirmed in a scoliosis case cohort. It may be possible that scoliosis develops secondary to other diseases that could also increase the risk of MACE. Future genetic analyses would be beneficial to assess causality through Mendelian randomisation techniques. Likewise, studies are required to determine whether the cardiovascular changes observed are reversible with scoliosis treatment surgery.

A previous study assessed 201 scoliosis patients for cardiopulmonary changes following corrective scoliosis surgeries and suggested that untreated scoliosis can result in pulmonary dysfunction and subsequently lead to right heart failure, increasing mortality[3]. Postoperative normalisation of cardiac measures was observed in this study, highlighting the potential benefits of surgical correction of scoliosis on cardiac function[3]. Additionally, a long-term follow-up study showed increased risk of pulmonary limitations such as shortness of breath in patients with untreated scoliosis, aggravated by spinal curvature[24].

It is challenging to accurately assess the cardiac function of scoliosis patients, due to the heart being displaced[22] and participants with scoliosis in the UKB had significantly less CMR scans available, possibly due to lack of comfort for prolonged periods of time in MRI machines. Adaptations may be required to allow patients with scoliosis to undergo MRI scans more comfortably, to further assess any alteration in cardiac function in scoliosis patients.

### Participants with scoliosis have altered cardiac diastolic strain rates

In the UKB, participants with scoliosis showed increased pressure at the top and bottom of the heart, as well as elongation of the sides of the heart. These results concur with the 2D-derived CMR findings of significantly increased PDSR_rr_ and decreased PDSR_ll_. A reduced PDSR_ll_ has been previously associated with reduced left atrial function[21]. No significant associations were found with the other studied CMR traits, suggesting that this compression does not alter blood flow through the heart. This altered cardiac strain may be explained by the deformity of the thoracic cage in scoliosis limiting cardiac diastolic movement. The mechanical abnormalities of the thoracic spine as well as the impact on the pulmonary system, may be the primary cause of the heart involvement[25,26]. Secondary involvement via altered pulmonary haemodynamics may be possible, where spinal curvature impacts pulmonary pressures leading to pulmonary hypertension. Likewise, direct compression of the myocardium could occur in conjunction to pulmonary involvement. These events can contribute to the development of cardiac consequences in scoliosis patients[27]. It would be beneficial to include cardiac follow-ups in patients with scoliosis to observe any cardiac alterations as scoliosis progresses, ensuring early intervention.

### Prevalence of scoliosis in the UK Biobank

We report a prevalence of scoliosis of 0.8% in the UKB population cohort. Previous literature has reported a scoliosis prevalence of 8% in adult volunteers aged over 40 years old[5]. Patients included in the previous clinical study were recruited on evaluation of bone mineral density and thus were likely at increased risk of scoliosis. Additionally, this discrepancy may be due to limitations of the UKB cohort (see *Limitations*).

Participants with scoliosis in the UKB are more elderly compared to the rest of the population, regardless of sex. Scoliosis occurs alongside other diseases such as osteoporosis or degenerative spine disorders with increasing age[5].

In the UKB cohort, scoliosis is more commonly found in females than males (2.5:1), which agrees with a previous scoliosis case cohort study of adolescents and adults that reported a ratio of scoliosis in females to males as 2:1[28]. Although not fully understood, there are different theories linking the higher prevalence of scoliosis in females, including implication of the autonomic nervous system (ANS) in skeletal growth and/or puberty through leptin hormone, which has been found decreased in female patients with idiopathic scoliosis[1,29,30]. The role of leptin in scoliosis is not fully understood, and analyses to determine causality would aid our current understanding of the mechanisms behind scoliosis. Furthermore, it has been reported that approximately 30% of females (n=324) suffer from systemic osteopenia which strongly contributes to the progression of spinal curvature in adolescent females following skeletal maturity[8].

### Limitations

The participants in the UKB cohort were recruited at 40-69 years of age and most participants are of European ancestry. Selection bias for inclusion in CMR sub cohort may have excluded more severe scoliosis participants from study. In addition, discrimination of scoliosis aetiology was mostly unspecified.

## Conclusions

This work describes for the first time in an adult population, evidence for altered cardiac function in adult participants with scoliosis. We identified altered diastolic strain, increased lifetime risk of MACE driven by HF and atrial fibrillation and observed cardiac compression in the UK Biobank participants with scoliosis. Further research is required to follow up the role of scoliosis in cardiac manifestations in a clinical setting.

## Data Availability

UK Biobank (https://www.ukbiobank.ac.uk/) population reference datasets are publicly available. All analysis code will be available on GitHub (https://github.com/ImperialCardioGenetics/Scoliosis)

## Competing interests

J.S.W. has consulted for MyoKardia, Inc., Foresite Labs, and Pfizer. D.P.O. has consulted for Bayer.

## Acknowledgements and funding sources

This work was supported by the Wellcome Trust [107469/Z/15/Z; 200990/A/16/Z], Medical Research Council (UK), British Heart Foundation [RG/19/6/34387, RE/18/4/34215], and the NIHR Imperial College Biomedical Research Centre. The views expressed in this work are those of the authors and not necessarily those of the funders. For open access, the authors have applied a CC BY public copyright license to any Author Accepted Manuscript version arising from this submission.

## CRediT statement

Conceptualization: K.A.M.; Methodology: K.A.M, V.Q.S.; Formal Analysis: V.Q.S., A.C.; Resources: J.S.W., D.O’R.; Data curation: V.Q.S., K.A.M.; Writing – original draft: V.Q.S.; Writing – review & editing: A.C., D.O’R., J.S.W., K.A.M.; Visualization: V.Q.S.; Supervision: K.A.M., J.S.W.

## Notes

### Competing Interest Statement

J.S.W. has consulted for MyoKardia, Inc., Foresite Labs, and Pfizer. D.P.O. has consulted for Bayer. All other authors declare no potential conflict of interest.

## References

1 Cheng JC, Castelein RM, Chu Winnie, et al. Adolescent idiopathic scoliosis. Nat Rev Dis Primers 2015;1:15030. doi:10.1038/nrdp.2015.30

2 Rummey C, Flynn JM, Corben LA, et al. Scoliosis in Friedreich’s ataxia: longitudinal characterization in a large heterogeneous cohort. Ann Clin Transl Neurol 2021;8:1239–50. doi:10.1002/acn3.51352

3 Sarwahi V, Galina J, Atlas A, et al. Scoliosis Surgery Normalizes Cardiac Function in Adolescent Idiopathic Scoliosis Patients. Spine (Phila Pa 1976) 2021;46:E1161–7. doi:10.1097/BRS.0000000000004060

4 Trobisch P, Suess O, Schwab F. Idiopathic Scoliosis. Dtsch Arztebl 2010;107:875–84. doi:10.3238/arztebl.2010.0875

5 Kebaish KM, Neubauer PR, Voros GD, et al. Scoliosis in adults aged forty years and older: Prevalence and relationship to age, race, and gender. Spine (Phila Pa 1976) 2011;36:731–6. doi:10.1097/BRS.0b013e3181e9f120

6 Aftzoglou P. Sarcopenia and falls in patients with adult scoliosis. J Frailty Sarcopenia Falls 2017;02:83–7. doi:10.22540/jfsf-02-083

7 Konieczny MR, Senyurt H, Krauspe R. Epidemiology of adolescent idiopathic scoliosis. J Child Orthop 2013;7:3–9. doi:10.1007/s11832-012-0457-4

8 Hung VWY, Qin L, Cheung CSK, et al. Osteopenia: A new prognostic factor of curve progression in adolescent idiopathic scoliosis. Journal of Bone and Joint Surgery 2005;87:2709–16. doi:10.2106/JBJS.D.02782

9 Bülbül S, Benlİ İT, Güler M, et al. Scoliosis prevalence in congenital heart disease patients. Turkish Spine 2013;24:3–12.

10 Kaito T, Shimada M, Ichikawa H, et al. Prevalence of and Predictive Factors for Scoliosis After Surgery for Congenital Heart Disease in the First Year of Life. Journal of Bone and Joint Surgery 2018;3:e0045. doi:10.2106/JBJS.OA.17.00045

11 Wang Y, Chen G, Xie L, et al. Mechanical factors play an important role in pectus excavatum with thoracic scoliosis. J Cardiothorac Surg 2012;7:2–4. doi:10.1186/1749-8090-7-118

12 Swank SM, Winter RB, Moe JH. Scoliosis and Cor Pulmonale. Spine (Phila Pa 1976) 1982;7:343–54. doi:10.1097/00007632-198207000-00004

13 Sudlow C, Gallacher J, Allen N, et al. UK Biobank: An Open Access Resource for Identifying the Causes of a Wide Range of Complex Diseases of Middle and Old Age. PLoS Med 2015;12:1–10. doi:10.1371/journal.pmed.1001779

14 Szustakowski JD, Balasubramanian S, Sasson A, et al. Advancing Human Genetics Research and Drug Discovery through Exome Sequencing of the UK Biobank. medRxiv Published Online First: 2020. doi:10.1101/2020.11.02.20222232

15 Bai W, Suzuki H, Huang J, et al. A population-based phenome-wide association study of cardiac and aortic structure and function. Nat Med 2020;26:1654–62. doi:10.1038/s41591-020-1009-y

16 Schafer S, de Marvao A, Adami E, et al. Titin-truncating variants affect heart function in disease cohorts and the general population. Nat Genet 2017;49:46– 53. doi:10.1038/ng.3719

17 Biffi C, de Marvao A, Attard MI, et al. Three-dimensional cardiovascular imaging-genetics: A mass univariate framework. Bioinformatics 2018;34:97–103. doi:10.1093/bioinformatics/btx552

18 Duan J, Ghalib B, Schlemper J, et al. Europe PMC Funders Group Automatic 3D bi-ventricular segmentation of cardiac images by a shape-refined multi-task deep learning approach. 2019;38:2151–64. doi:10.1109/TMI.2019.2894322.Automatic

19 Meyer H v, Dawes TJW, Serrani M, et al. Europe PMC Funders Group Genetic and functional insights into the fractal structure of the heart. Nature 2021;584:589–94. doi:10.1038/s41586-020-2635-8.Genetic

20 McGurk KA, Zheng SL, Henry A, et al. Correspondence on “ACMG SF v3.0 list for reporting of secondary findings in clinical exome and genome sequencing: a policy statement of the American College of Medical Genetics and Genomics (ACMG)” by Miller et al. Genetics in Medicine 2022;24:744–6. doi:10.1016/j.gim.2021.10.020

21 Thanaj M, Mielke J, McGurk KA, et al. Genetic and environmental determinants of diastolic heart function. Nature Cardiovascular Research 2022;1:361–71. doi:10.1038/s44161-022-00048-2

22 Li S, Yang J, Zhu L, et al. Ventricular and atrial mechanics and their interaction in patients with congenital scoliosis without clinical heart failure. Cardiol Young 2014;760:976–83. doi:10.1017/S1047951114001504

23 Mauro A lo, Aliverti A. Physiology of respiratory disturbances in muscular dystrophies. Breathe 2016;12:318–27. doi:10.1183/20734735.012716

24 Hawes MC, Weinstein SL. Health and Function of Patients with Untreated Idiopathic Scoliosis. J Am Med Assoc 2003;289:2644–5. doi:10.1001/jama.289.20.2644-a

25 Janicki JA, Frcsc BA. Scoliosis: Review of diagnosis and treatment. Paedriatric Child Health 2007;12:771–6.

26 Prof A. Pulmonary Functions in Patients With Idopathic Scoliosis. 2017;90:123–8.

27 Li Q, Zeng F, Chen T, et al. Management of Severe Scoliosis with Pulmonary Arterial Hypertension: A Single-Center Retrospective Case Series Study. Geriatr Orthop Surg Rehabil 2022;13:1–11. doi:10.1177/21514593221080279

28 Lang C, Wang R, Chen Z, et al. Incidence and Risk Factors of Cardiac Abnormalities in Patients with Idiopathic Scoliosis. World Neurosurg 2019;125:e824–8. doi:10.1016/j.wneu.2019.01.177

29 Man GCW, Tam EMS, Wong YS, et al. Abnormal Osteoblastic Response to Leptin in Patients with Adolescent Idiopathic Scoliosis. Sci Rep 2019;9:1–7. doi:10.1038/s41598-019-53757-3

30 Wang Q, Wang C, Hu W, et al. Disordered leptin and ghrelin bioactivity in adolescent idiopathic scoliosis (AIS): a systematic review and meta-analysis. J Orthop Surg Res 2020;15:1–9. doi:10.1186/s13018-020-01988-w

